# Zeroth-Order Deterministic Local and Delay Pandemic Model Comparison

**DOI:** 10.1101/2020.08.05.20162164

**Authors:** David L. Bartley

**Affiliations:** Centers for Disease Control and Prevention, Retired, 3904 Pocahontas Avenue, Cincinnati, Ohio 45227

**Keywords:** pandemic, models, delay, functional, retarded, Covid-19

## Abstract

Delay differential equations are set up for zeroth-order pandemic models in analogy with traditional SIR and SEIR models by specifying individual times of incubation and infectiousness prior to recovery. Independent linear delay relations in addition to a nonlinear delay differential equation are found for characterizing time-dependent compartmental populations. Asymptotic behavior allows a link between parameters of the delay and traditional models for their comparison. In analogy with transformation of the traditional equations into linear form giving populations and time in parametric form, expansion in the delay provides a simple recursive solution. Also, a soliton-like solution in terms of a logistic function can be applied for accurate approximation. Otherwise, straightforward numerical solution is effected in terms of linearized boundary conditions specifying the distribution of instigators as to their initial infection progress—in contrast to traditional models specifying only initial average infectious and exposed populations. Examples contrasting asymptotically-linked traditional and delay models are given.

## Introduction

The Covid-19 epidemic has aroused great interest in pandemic modelling. This paper addresses two types of compartmental deterministic models. Such models compute the evolution of various populational *compartments* following initial infection Considered here are the classic SIR [7] and SEIR Models which address Susceptible, Exposed, Infectious, and Recovered (including deceased) populations. For a summary and details of these models and their many variations, see [8] and references within. Most [2, 6-9] of these models are expressed in terms of ordinary nonlinear differential equations local in the time.

Less common are models [1, 5, 10-12, 14, 15] which specify the rates of change in the populations at a given time in terms of populations at an earlier time. Specifically, this paper considers a pandemic model similar to the traditional SEIR Model (including SIR) constructed by specifying the incubation time *τ_δ_* (for example, 5 days) an individual spends prior to becoming infectious and the total time *τ_γ_* (e.g., 20 days) to recovery. The traditional instantaneous differential equations of the SEIR Model are replaced by *delay* or *functional* nonlocal nonlinear differential equations [3] solvable numerically [13] in terms of the prior history of the initially exposed or infectious. Since in reality, both *τ_δ_* and *τ_γ_* may vary widely between individuals, this type of model, together with the instantaneous or local models, must be considered 0-th order approximations.

The plan for this paper is as follows. First, the delay differential equations are presented, together with various simple relationships among the populations. Asymptotic limits are given. Comparison of the instantaneous and delay models is possible by linking the parameters of the two types of models together requiring asymptotic coincidence. Boundary conditions for the delay models are established that permit specifying the initial distribution of the pandemic instigators in terms of their initial disease progress, unlike the instantaneous models. The models are also comparable through the finding of accurate solutions to the delay SIR model in analogy to solution of the usual instantaneous model with populations and time in parametric form from equations transformed into linear form, obviating addressing the differential equations numerically. Finally, several numerical comparisons are made.

## Equations of Motion

As with the instantaneous SEIR Model, the rate of new infections per unit time is approximated by:

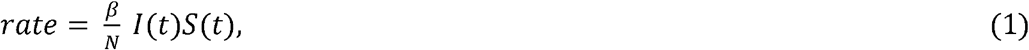

where *β* is a rate constant, I is the number of infectious individuals at t, S is the number of those susceptible to infection, and N is the (constant) total number of individuals. Following exposure, suppose that an incubation time *τ_δ_* is required until an individual becomes infectious and that a further time interval *τ_γ_ − τ_δ_* remains until the individual either recovers or dies.

Suppose the pandemic begins at *t* = 0 with the introduction of a small number of exposed or infectious individuals. By the time *t* = τ*x_γ_*, all the instigators will have recovered (or died), after which the following conditions hold. The S susceptible individuals at time *t* > *τ_γ_* consist in all present (*N − E*(0) − 1(0)) at *t* = 0+ minus those who have become infected after *t* = 0. Therefore,

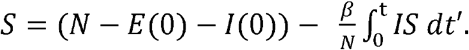

At time t, the E exposed individuals are all who suffered new exposures between *t* − *τ_δ_* and *t*. Therefore,

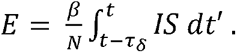

Similarly, the I infectious individuals at t (> *τ_γ_*) consist exactly in those who suffered new exposures between *t* − *τ_γ_* and *t* − *τ_δ_*. Therefore,

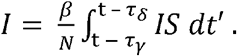

At *t* > *τ_γ_*, the R recovered or deceased individuals at t are those who suffered new exposures before *t* − *τ_γ_* plus the (*E*(0) + I(0)) initially exposed or infectious. Therefore,

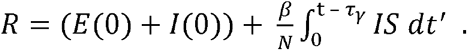

The above equations imply, of course, that:

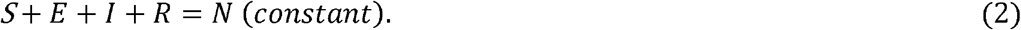

Differentiating the above equations with respect to time t results in an equivalent set of non-local non-linear differential equations:

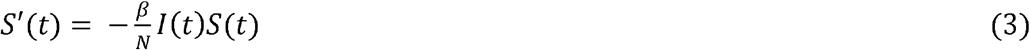

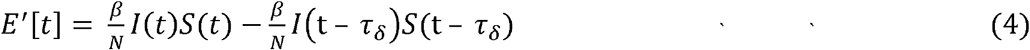

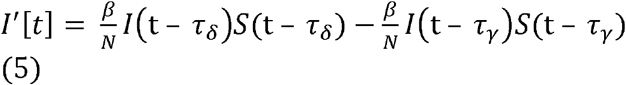

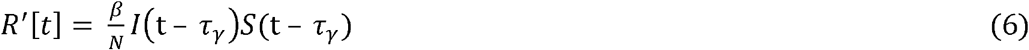

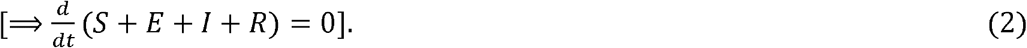

The equations are valid for *t* > *τ_γ_* and depend for solution on the *functions* I(t) and S(t) given over the interval, 0 < *t* < *τ_γ_* as the initial values, unlike the boundary conditions at a *single point with* ordinary differential equations, hence the aptness of the term, *functional* differential equation. As detailed below, the boundary conditions, I(t) and S(t) over this early interval, are determined by the *distribution* in the exposure times of the initially exposed or infected who trigger the pandemic. Equations (3-6) specialized to a Delay SIR model are identical to the model discussed in References [11, 12, 15], neglecting natural birth and death.

The above equations are analogous to the local SEIR Model in widespread use:

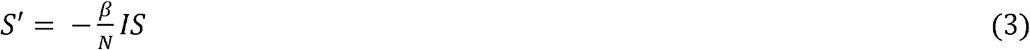

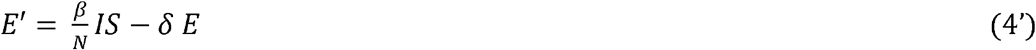

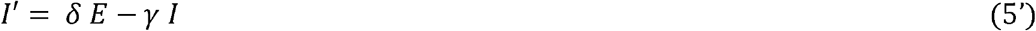

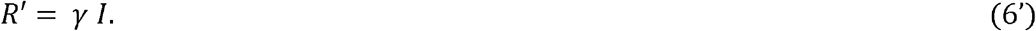

### Relationships

Both the delay and instantaneous models are characterized as above by actually only *three* equations in the three functions S, E, and I. In fact, there is a simple relation between S and R.

### SEIR-

In the case of the instantaneous model, Equations (1 and 4’) imply that

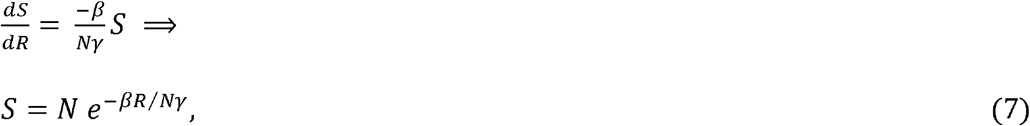

which satisfies S(0) ~ N at R = 0 (at t = 0).

### Delay SEIR

With the less familiar delay model, Equation (7) holds only asymptotically (at t → ∞ or 0). In addition to

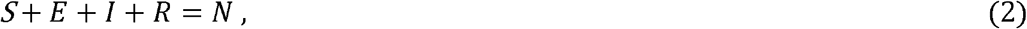

there are two independent nonlocal relations among S, E, I, and R. At t > *τ_γ_*, new infections created at t result in new recoveries at t + *τ_γ_*, so the recovery rate must lag behind the creation rate by *τ_γ_*. Explicitly, Equations (3 and 6) imply that

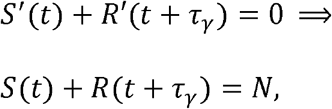

where the integration constant Ms determined by the *t* → ∞ limit. Equations (4 and 5) then imply:

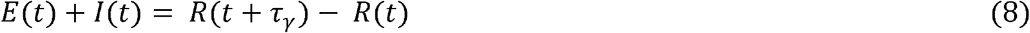

Similarly, infectiousness lags infection by *τ_δ_*, and Equations (3 and 4) give:

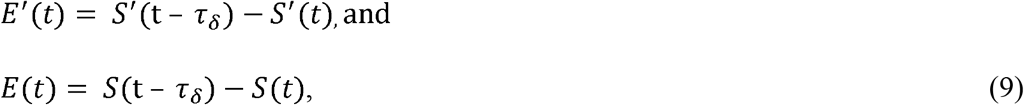

as the *t* → ∞ limit ⇒ the integration constant = 0. As easily seen, the above relations hold for the simpler SIR Models as well (setting E and *τ_δ_* equal to zero). Equations (2, 3, 8, and 9) together with boundary conditions are then sufficient to determine the evolution of the four populations.

### Asymptotic limits

#### SEIR

Another similarity between the delay and instantaneous model is found in the asymptotic limits (*t* → ∞) of S and R. As *t* → ∞, I and E → 0, and so

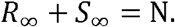

In the familiar case of the local SEIR model [6, 9], Equation (7) implies:

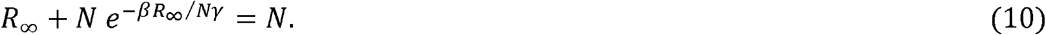

Solutions *R_∞_* of this transcendental equation have been tabulated as the Lambert W function or the ProductLog in *Mathematica*.

### Delay SEIR

The Delay SEIR Model is somewhat more complicated, but results in an expression of the same form as Equation (10). One approach is to expand the various functions in *τ_γ_* and *τ_δ_*. Equation (8) implies:

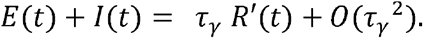

Equations (9 and 6) imply:

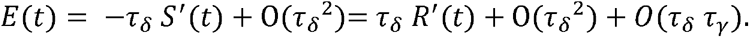

Therefore, keeping only terms linear in *τ_δ_* or *τ_γ_*,

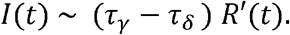

Equations (3) then gives

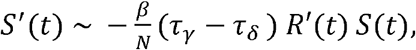

which integrates to:

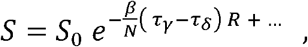

where *S*_0_ ensures *S*(*R*) = *S*_0_ at R = 0. Finally, in the limit t → ∞, where I and E → 0,

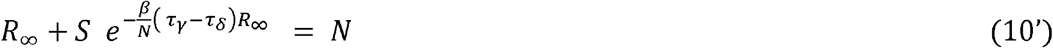

Again, the asymptotic value *R_∞_* is given in terms of the Lambert W function. *S_∞_* = *N − R*_∞_, since *I* → 0 *as t* → 1.

### Asymptotic linking of the models

In the sections below, the delay and instantaneous models are placed on an equal footing for comparison by identifying the infectious-time constant *γ*^-1^ with (*τ_γ_* − *τ_δ_*), the time an individual is infectious, and the exposed-time constant *δ*^−1^ with *τ_δ_*, i.e., with the means of the corresponding exponential distributions. Also, as indicated by Equations (10 and 10’), the former identification forces the asymptotic limits to be identical.

## Linearized Boundary Conditions

Although the above delay differential equations are non-linear, near the start of a pandemic with a small number of infected individuals, a linearized version of the equations is extremely accurate. The corresponding solutions for t < *τ_γ_* are simple and transparent. Furthermore, superposition makes possible the combination of the initially infected as distributed over a range of infection stages. Therefore, the boundary conditions (for time t < *τ_γ_*) that reflect the history of the initially infected are easily determined and permit solution of the nonlinear delay equations at t > *τ_γ_* using the same numerical methods as with instantaneous nonlinear equations.

### Delay SIR Model

The earliest times in a pandemic are special in that only the initially infected can recover at t < *τ_γ_*, within which explicit solution of s[t] is possible. For example, with the Delayed SIR Model, as long as no recovery occurs by time t,

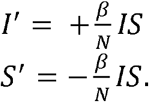

These equations are easily solved in terms of the logistic function, since S + I + R = N (and R = 0 and N are constants). However, for the reasons outlined above, at t < *τ_γ_* the linearized equation is adopted:

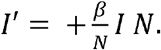

Then an initially infectious component *I*_0_ with time t_r_ remaining prior to recover results in infections *I*(*t*) expanding exponentially in time as:

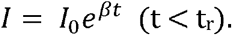

Then following recovery of the initial infection at t = t_r_,

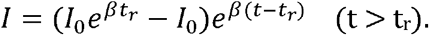

In other words,

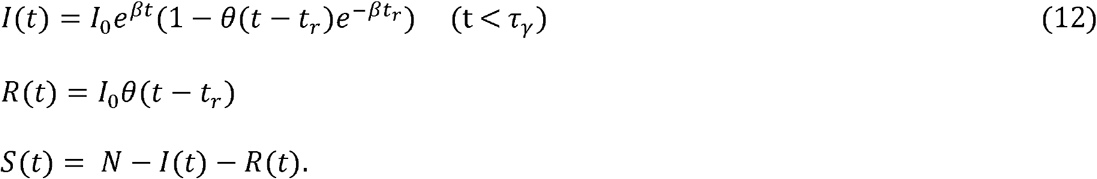

Now if there is a distribution *ρ*(*t_r_*) in the time *t_r_* to recovery of the initially infectious, then superposition implies that the infectious *I*, recovered *R*, and susceptible *S* at early times are given by

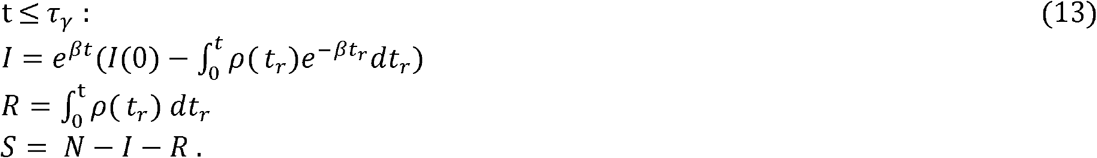

These easily-coded functions of t supply the required information for numerical solution of the nonlinear delay differential equations for later time t. The remarkable feature of the delay model is that solutions I(t), R(t), and S(t) depend on an entire initial function, *ρ*(*t_r_*).

Incidentally, the density *ρ*(*τ_i_*, *t*) at later times (t > 2*τ_γ_*) of the infectious vs the length of time *τ_i_* infectious (i.e, *τ_γ_ − τ_i_* = t_r_ is the time to recovery) is given by:

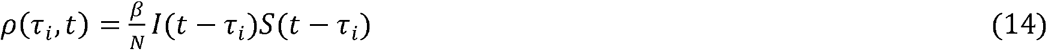

which is justified by:

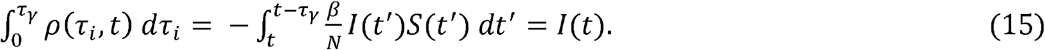

Any “old” case prior to *t* − *τ_γ_* has “recovered” by the time t; Equation (15) expresses the fact that only the new infections need to be considered for the density at time t. The boundary conditions for the delayed SEIR Model can be expressed similarly to Equations (13), but are somewhat more complicated and are often more simply determined by numerical solution.

## Partial Solutions

### SIR

The traditional SIR Model is easily transformed [4] into a set of linear equations by replacing the independent variable *t* by an alternative variable *η* via *I*(*t*)*dt* = *dη*. Then *I*(*η*) and *S*(*η*) can be easily expressed in closed form, and *t*(*η*) is determined by evaluating the integral ∫ *I*(*η*)^−1^*dη* numerically. The resulting parametric representation of I, S, and t in terms of *η* replaces numerical solution of the original nonlinear differential equations.

### Delay SIR

No similar transform of the Delay SIR Model is known. One approach found accurate results from expansion in the delay (Appendix A). Alternatively, numerical experimenting indicates that varying the boundary conditions at the start of a pandemic, holding *β* and *τ_γ_* constant, very accurately simply shifts the compartment curves in time (at the scale of the figures in this paper). This indicates the possibility of a soliton-like solution extending from t = −∞ to +∞. Furthermore, the symmetry and shape of the curves depicting the infectious population and the new-infection rate suggest a logistic rather than gaussian model, at least in approximation. Therefore, a soliton-like susceptible population may be taken tentatively as:

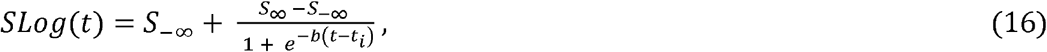

where *S_∞_* represents the asymptotic value given in Equation (10’) in terms of *S*_0_, and *βτ_γ_*, and *t_i_* is the time of inflection in the susceptible population. The parameters *S*_−∞_ and b are determined to best solve the equations of motion (3 and 8’) for the susceptible population.

Equation (8) can be rewritten as:

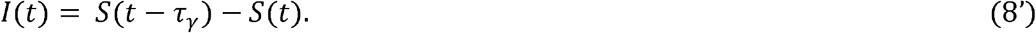

Equations (3 and 8’) then imply:

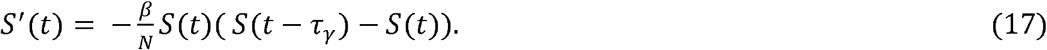

Substituting Slog(t) for S(t), results in an exact solution of Equation (17), iff

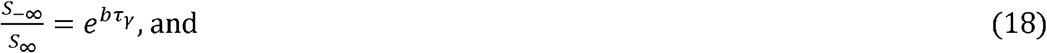

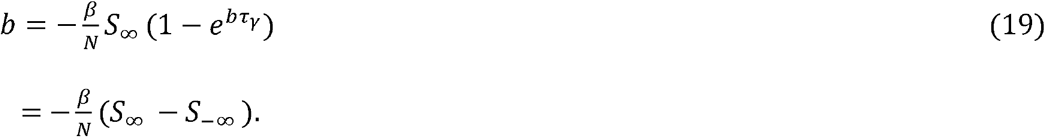

Equation (19) is a transcendental equation for *b* with evaluation in Mathematica as:

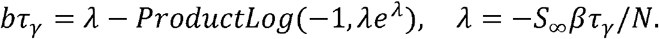

For an independent complementary derivation of this result, see Reference [11].

The remaining parameter *t_i_* is arbitrary, and so any solution Slog(t) translated in time is also a solution of (17) with the identical shape. Though the solution Slog(t) does not satisfy boundary conditions as spelled out above, a value for the inflection time *t_i_* can be easily found so that Slog(*τ_γ_*) matches the boundary value *S*(*τ_γ_*). The result, though still not satisfying the boundary conditions over tin [0, *τ_γ_]* except at the end point *τ_γ_*, results in a remarkably accurate solution as compared to the exact *S*(*t*) at large *t* as found numerically [13].

For example, Fig. 1 shows the error in such an approximation for three values of the time *t*_r_ to recovery of the pandemic instigators (using parameter values as in the following section).

**Figure 1.**
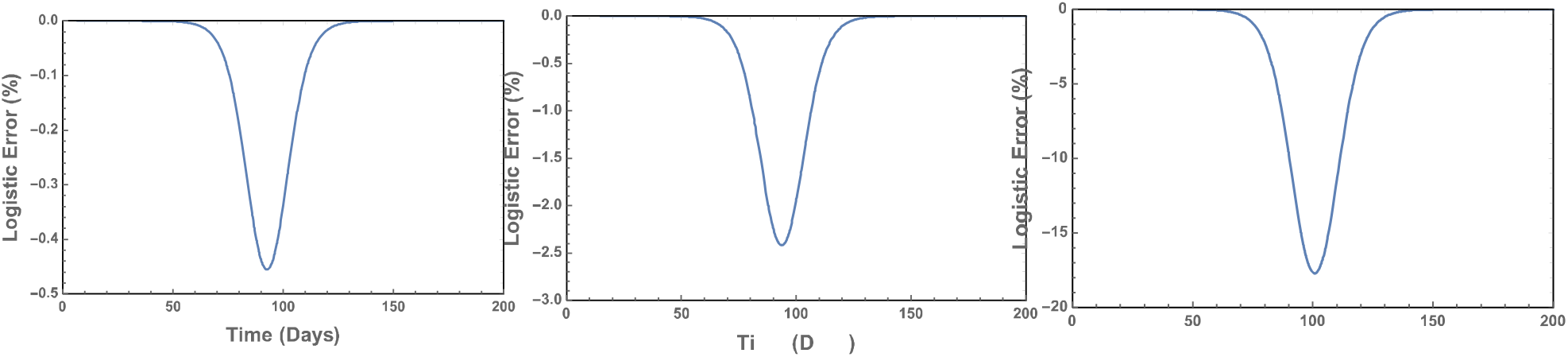
Error of the logistic approximation relative to the exact susceptible population for the time *t*_r_ to recovery of the instigators equal to 15 days, 7.5 days, and 1 day (left to right).

The error is (excessively) exaggerated near the inflection in the susceptible population. For example, in the worst-case situation with *t*_r_ - equal to 1 day, global agreement between approximation and exact curve is remarkable (see Fig. 2).

**Figure 2.**
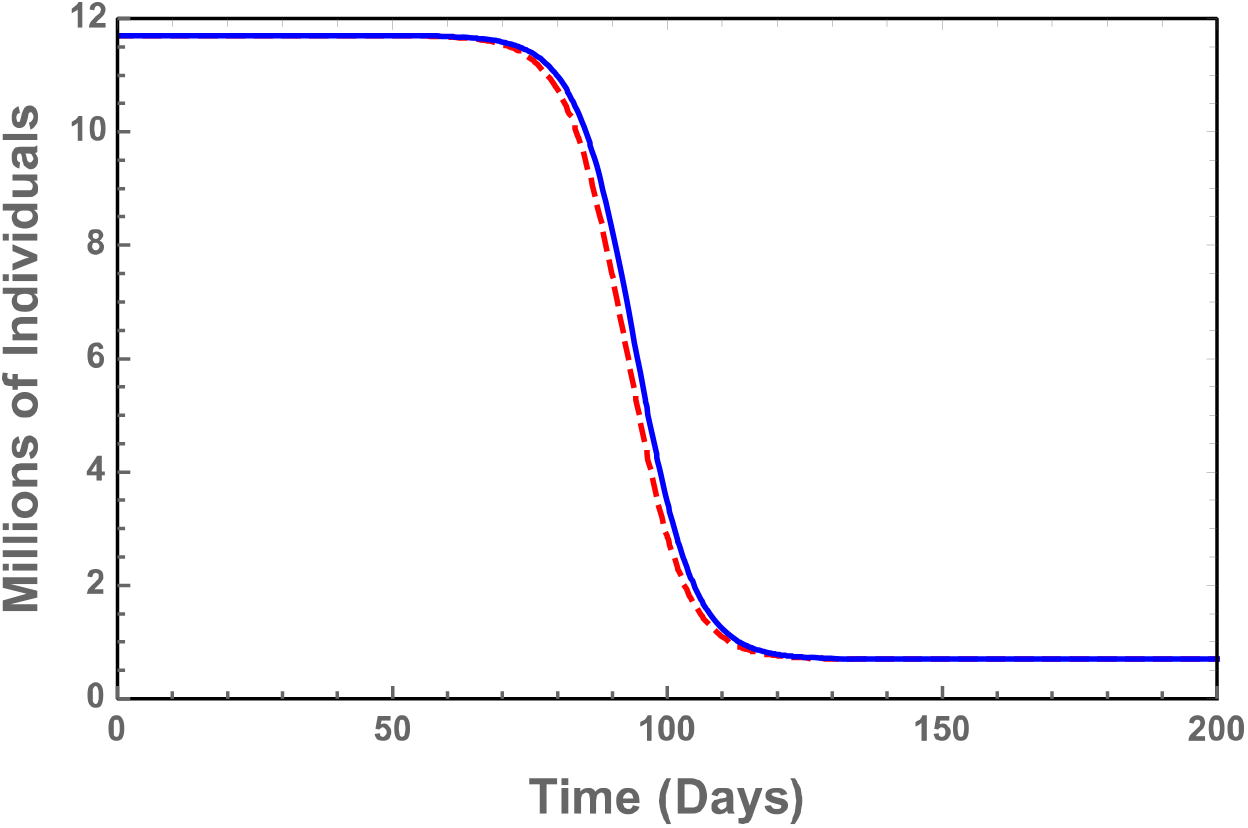
Agreement between logistic approximation (dashed) and exact (solid) susceptible populations at the time *t*_r_ o recovery of the pandemic instigators taken to equal 1 day.

The close agreement seen here is partly because not only are *S*(*τ_γ_*) and Slog(*τ_γ_*) exactly matched, but also the 1^st^ derivatives are close as well. At *t* ~ *τ_γ_*, S(t) ~ *N* − *I*_0_*e*^_^*^βt^*, while Slog(t) ~ *N − I*_0_*e^−bt^*, and b ~ *β*. Matching Slog(t) to the boundary function approximates matching the solution to a 2^nd^ degree ODE at the given boundary. Perhaps the most useful result of this exercise is the realization that the large-t functions S, I, and R lose much of the detail originally in the boundary interval [0, *τ_γ_*].

## Numerical Comparisons

The differences between the delay and local models are best illustrated by means of numerical examples. Parameters for the above models were selected as follows:

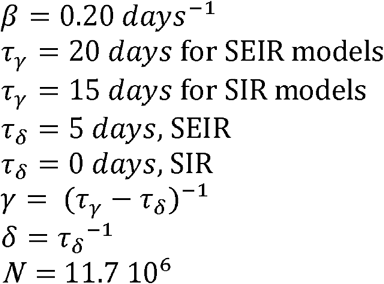

These parameters imply the infectiousicity index R_t_ is equal to 3 at t = 0 (i.e., at the start of the pandemic). Rt is the mean number of infections caused by an individual infectious from time t and can be computed in terms of the susceptible population S(t) as:

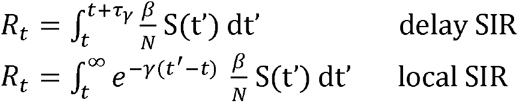

*R*_0_ and *R_∞_* are given simply in terms of S(0) and S(∞), in regions of t where S(t) hardly varies.

Illustrating Equations (13), an even initial exposure density *ρ*(*t_r_*) = 1/*τ_γ_* was selected for the Delay SIR Model and instigating population = 1 for the corresponding Local SIR Model. The integral in Equation (13) then easily provides sufficient information for numerical solution (using Mathematica) for arbitrary time. The results (assuming no mitigation) are illustrated in Figs. 3 and 4.

**Figure 3.**
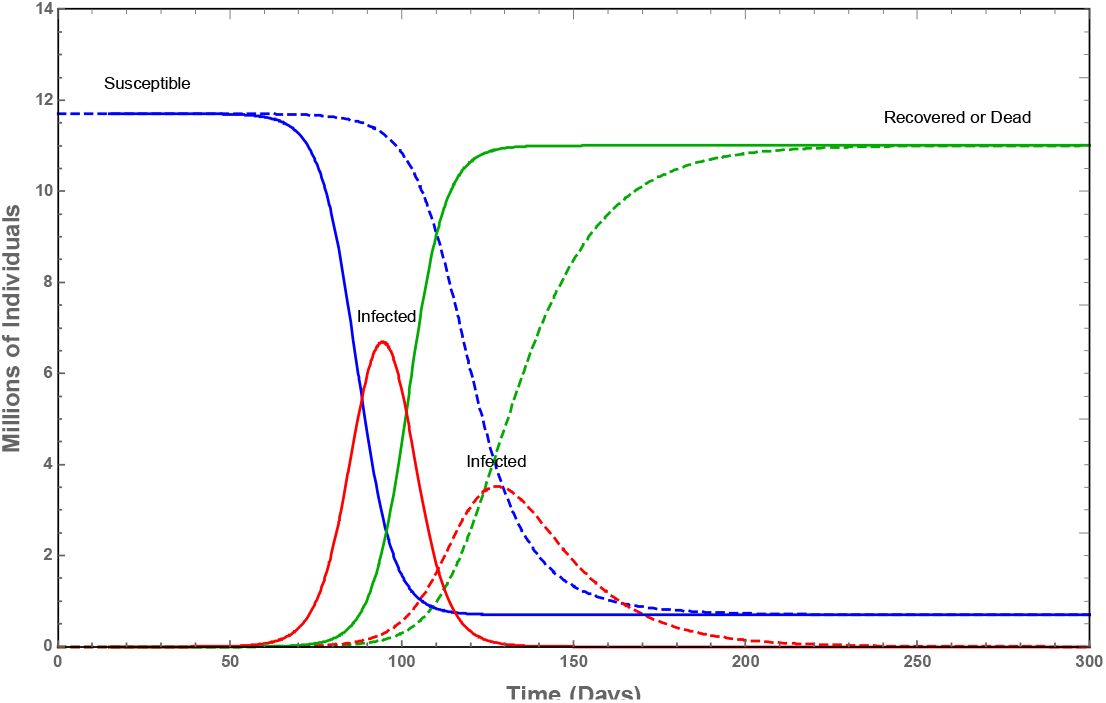
Comparison of the susceptible, infectious, and recovered (or deceased) populations from asymptotically identical SIR models, local (dashed) and delay (solid) with constant distribution of initial exposing factor at infected population = 1 (other parameters in text).

**Figure 4.**
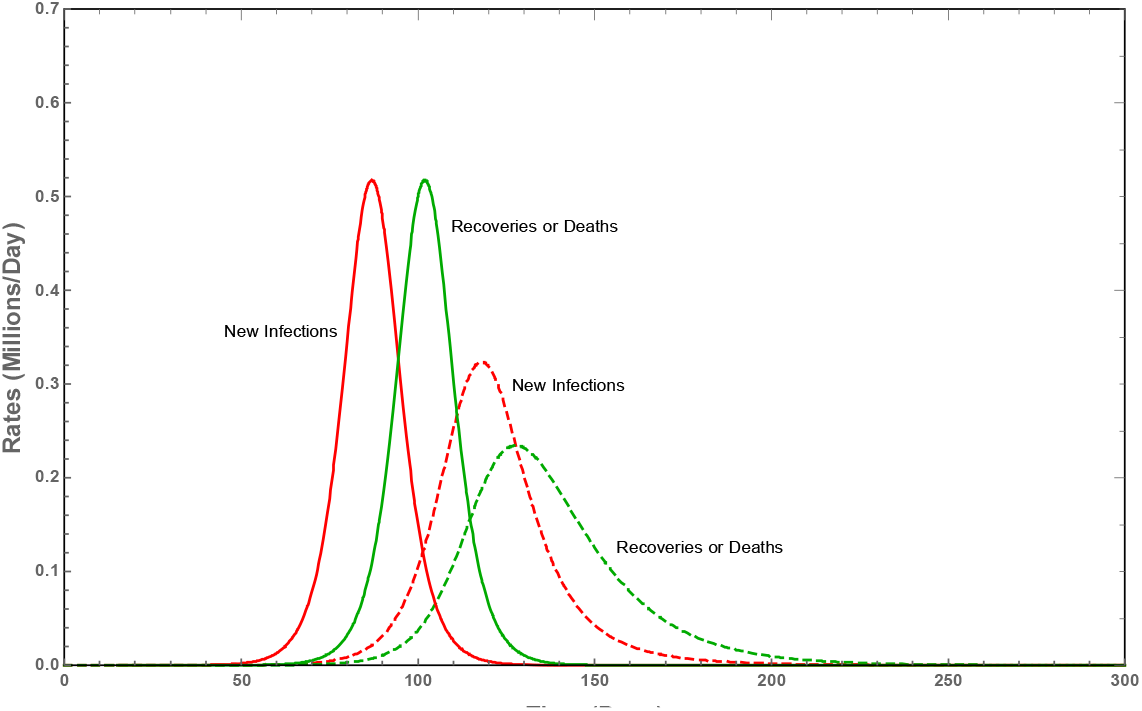
Comparison of the new infections/day and recoveries/day according to local (dashed) and (solid) delay SIR models.

Note the identical asymptotic behavior of the populations in accordance with Equations (10 and 10’). Also, the local curves lag behind those of the delay model, by 1-2 months. The curves for the new cases/day attain maxima at 87 days and 118 days for the delay model and local model, respectively. This lag is not surprising as the probability of continued infectiousness of an individual falls off as 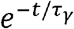 for the local model, whereas the probability remains equal to 1 from t = 0 until recovery at t = *τ_γ_*. Also, note the near-symmetry of the curves from the delay model and close relation to the logistic function.

At the opposite extreme from an even density, calculations were also done for sharply peaked densities at a variety of initial recovery times of the instigating individuals. The results for the SIR models are shown in Fig. 5. The time required for attaining a maximum (as in Fig. 4) in the new cases/day was determined for each initial recovery time. The curve for the delay model is remarkably flat, despite upward turning for the nearly recovered instigator, and, of course, the approach to infinity in the limit of an initially recovered individual. The difference between local and delay models is close to that of the even distribution over the initial recovery times, despite the nonlinearity of the equations.

**Figure 5.**
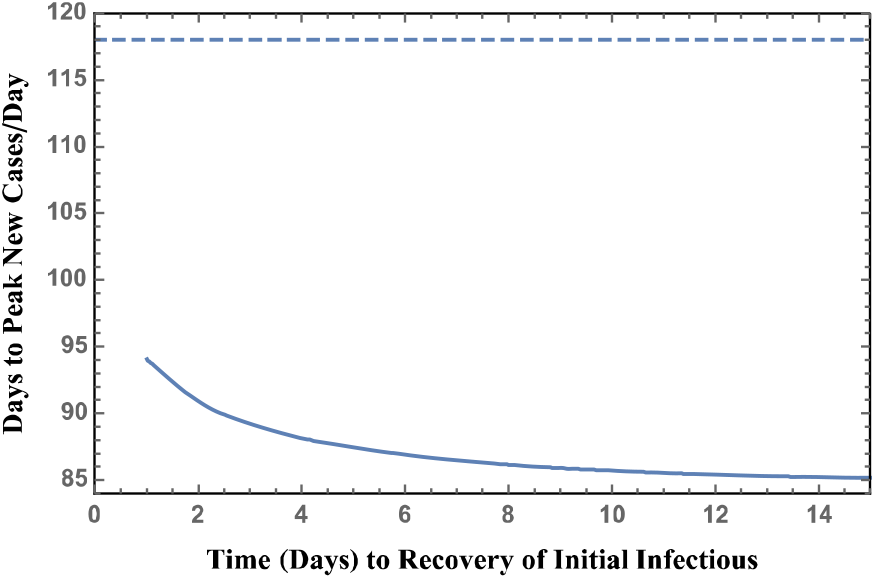
SIR: time to maximum new cases/day in terms of state of health of the initial exposing population = 1 at t = 0.

A similar calculation was done using the SEIR models. A typical result is shown in Fig. 6.

**Figure 6.**
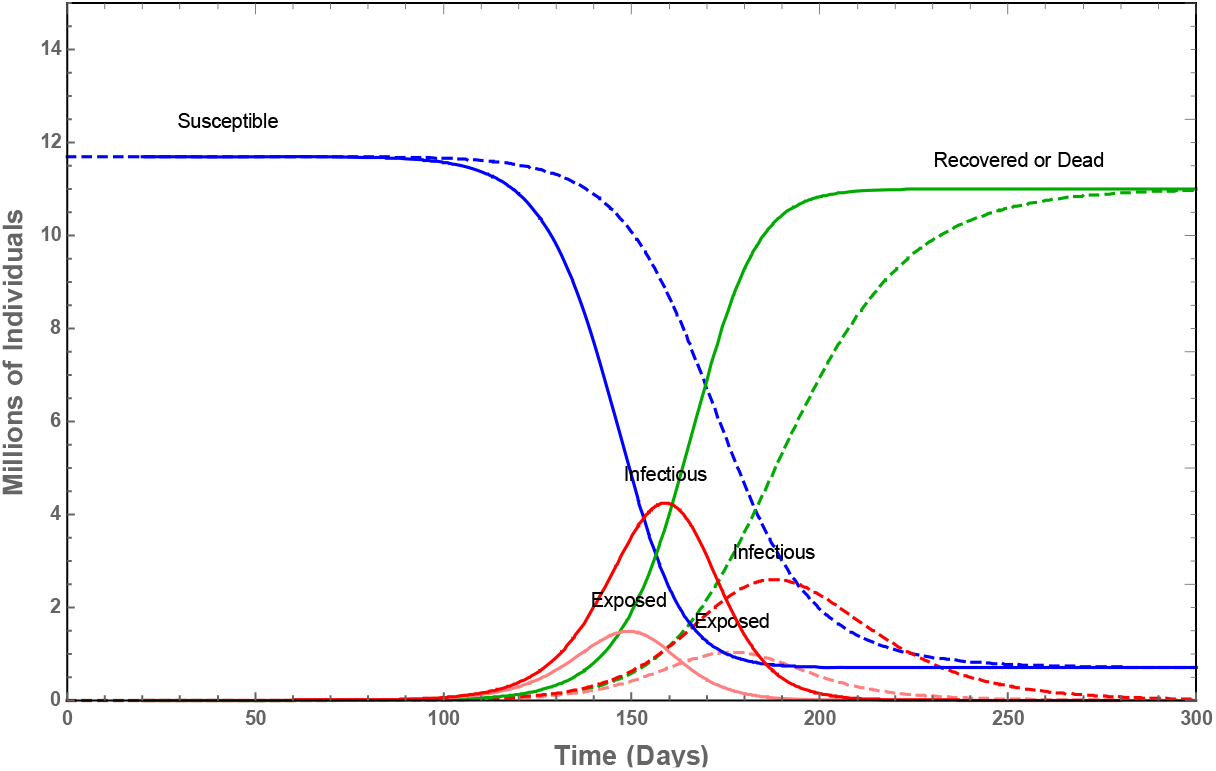
Typical solution for inclusion in the compilation of Figure 7. Local (dashed) and (solid) delay SEIR models.

**Figure 7.**
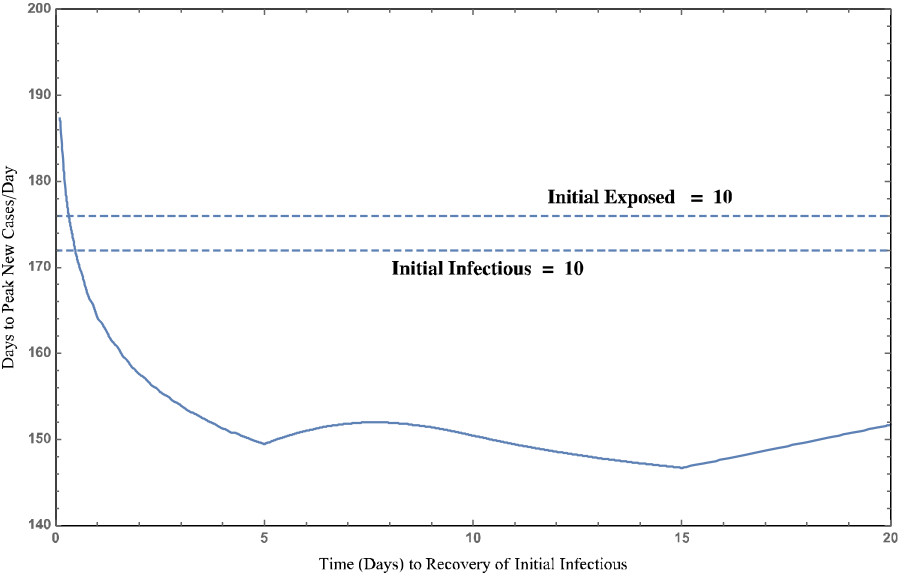
SEIR: time to maximum new cases/day in terms of state of health of the initial exposing population = 10 at t = 0 (other parameters in text). Dashed: local, Solid: delay.

In this case, the linearized boundary conditions were determined numerically. This required attention to the three separate situations: *t*_r_ *< τ_δ_, τ_δ_ < t*_r_ *< τ_γ_ − τ_δ_*, and *τ_γ_ − τ_δ_ < t*_r_ *< τ_γ_* where the initial instigators were only exposed prior to becoming infectious. Again, the increase in the time to maximum in the new cases/day on approaching nearly recovered instigators is apparent, yet is limited to the lowest few days prior to recovery. The difference between local and delay models is similar to that of Fig. 5 for the SIR model, although time from pandemic start to maximum in the new cases/day is naturally longer for the SEIR model with time required for infectiousness to begin.

## Conclusions

Similarities and differences are found between basic simple deterministic pandemic models—instantaneous vs delay. Both types of models expressed in terms of differential equations are readily addressed using established techniques of numerical analysis. Interestingly, though nonlinear, both can be expressed in terms of accurate closed-form solutions. Just as the instantaneous equations are very simply linearized in terms of population compartments and time in parametric form, the delay equations admit a simply-evaluated solution in terms of a soliton-like logistic function. Further research into this solution is merited, for example, determining how to extract relevant information contained in the initial boundary interval for more general models.

Both model types share the functional form of asymptotic values relevant to the pandemic winding down. This allows linkage between model types for comparison. Equivalently, parameters can be chosen to equate growth at the pandemic start.

A difference between the models exists in the form of the initial boundary conditions. The delay equations depend on an initial *function*. This function can be expressed in terms of the distribution of pandemic instigators as to initial disease progress. This expression is facilitated by adopting accurate linear boundary conditions permitting superposition even though the equations valid during the progress of the pandemic are nonlinear.

The results of this work show significant difference between the usual local and the delay models *between the* start and death of the pandemic. The difference no doubt relates to the individual’s probability of remaining infectious. With the usual local model, this probability falls rapidly as 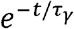 or 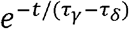 where t is the time from the start of infectiousness. In contrast, with the delay model considered here, the probability of infectiousness lasting until time t remains high at 1.00 until the time *τ_γ_* or *τ_γ_* − *τ_δ_* is reached.

This difference may be significant to the progress of a pandemic. The ultimate result as the pandemic winds down is identical for the two types of models considered here. Of course, it is the progress up to the time of the maximum in the new cases/day that is necessary to understand in order to adopt appropriate mitigating measures.

These results may provide help in choosing between the models. From the point of view of calculation, the delay model is not significantly more difficult to analyze. Which model is appropriate depends on the details of recovery or death of the individual following infection.

## Data Availability

No data are referred to.

## Appendix A – Solution of Delay SIR by expansion in the delay

In more complicated cases than treated in the text, a particular solution such as Equation (16) may be impractical to determine for approximation to the delay differential equations. However, another approach may be useful in some contexts. The following results in a partial solution. Equation (8) can be rewritten as:

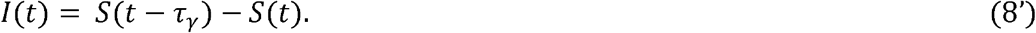

Expansion in 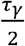 at 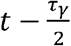 results in:

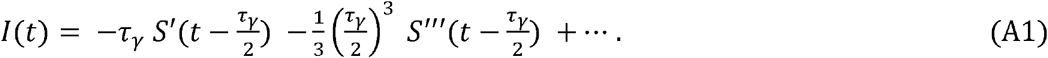

Keeping only the first term, Equation (3) simplifies to:

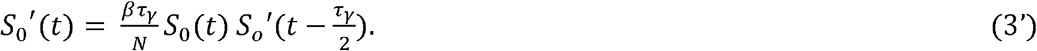

Equation (3’) is a *neutral* delay differential equation (i.e., with delay in the derivative [3]) which integrates to

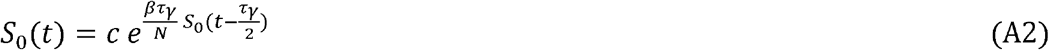

where the constant *c* is determined by matching at t = *τ_γ_*.

Equation (A2) is a recurrence relation ending after a finite number of iterations from any time *t > τ_γ_* down to within a boundary interval [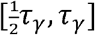, *τ_γ_*], where the initial *S*(*t*) is defined in terms of the pandemic instigators as described in the text above. Equation (A2) is evaluated for *S*_0_(*t*) automatically in programs such as *Mathematica*.

Numerical experimenting indicates that the global approximation *S*_0_(*t*) has an 0(*τ_γ_*) shift from S(t) near the peak in I(t) and inflection in S(t). This shift arises from the inaccuracy of the approximation (3’). Accounting for the 2^nd^ term in Equation (A1) results in an expression similar to Equation (A2). This allows a perturbative correction which “explains” most of this shift.

In any case, suppose S(t) is approximated (at *t* ≫ *τ_γ_*) as:

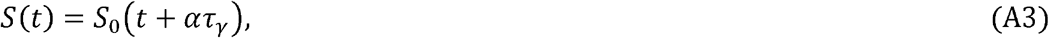

where *α* = *O*(1). The infectious population *I*(*t*) is then determined directly from Equation (8’). The accuracy of Equation (A3) (away from the boundary conditions near t = 0, yet retaining their causative effect) is illustrated in Fig. A1 by comparison to the exact numerical solution of the original nonlinear equations for two values of the infection rate *β*, with *α* = 1.25. Aside from *β*, the assumed conditions are as in Fig. 3. The fractional shift *α* depends weakly on the specifics of the boundary conditions, but is insensitive to *β* or *τ_γ_*. The extreme accuracy as to curve shape of this approximation with R_0_ between 2 and 3 and with *τ_γ_* at least as large as 30 is as yet not understood.

**Figure A1.**
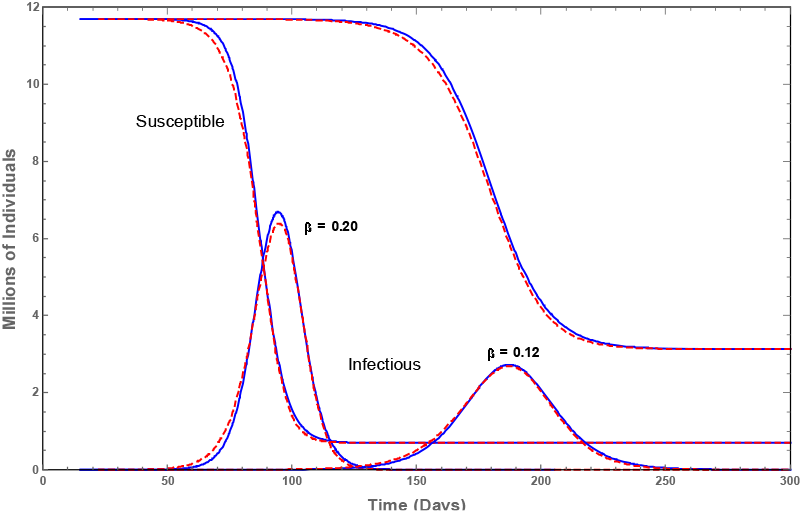
Comparison of approximate recursive (dashed) and exact numerical (solid) solutions of the delay SIR equations at *τ_γ_* =15 days for two different values of the infection rate *β*.

## Notes

### Competing Interest Statement

The authors have declared no competing interest.

### Funding Statement

No funding was involved in this research.

### Author Declarations

No health data are contained in the article.

### Summary of Updates

Improved Appendix A: Delay DE is expressed as infinite series in the delay, resulting in neutral delay equation solvable in approximation with likely application in some contexts.

